# Predictors of symptom trajectory in newly diagnosed ulcerative colitis: a 3-year follow-up cohort study

**DOI:** 10.1101/2023.11.29.23299204

**Authors:** Maaike Van Den Houte, Livia Guadagnoli, Lena Ohman, Magnus Simren, Hans Strid, Lukas Van Oudenhove, Jan Svedlund

## Abstract

**Introduction:** Psychological symptoms are associated with poorer ulcerative colitis (UC)-related outcomes. However, the majority of research to date is cross-sectional and longitudinal data is lacking. We aimed to identify patient subgroups based on the longitudinal evolution of GI symptom levels and health-related quality of life (HRQoL), and to disentangle the directionality of effects between GI symptom levels and levels of psychological distress.

**Methods:** Self-reported GI symptom severity, HRQoL, inflammatory biomarkers and psychological distress were assessed in 98 newly diagnosed UC patients at disease onset and yearly for 3 consecutive years. Latent class growth analysis (LCGA) was used to determine subgroups based on longitudinal trajectories of symptom severity (diarrhea, abdominal pain) and physical and mental HRQoL, and baseline predictors of trajectory group membership were determined. Cross-lagged structural equation models were used to disentangle temporal relationships between psychological functioning and GI symptom severity.

**Results:** Different subgroups were found based on the evolution of diarrhea, abdominal pain, and physical and mental HRQoL over time. Patients with higher initial psychological distress had increased probability of maintaining higher levels of diarrhea (p = 0.049) and abdominal pain (p = 0.009). Conversely, patients with lower initial levels of diarrhea (p < 0.001) and abdominal pain (p = 0.003) had higher chances of maintaining lower levels of psychological distress. Higher levels of C-reactive protein at baseline predicted greater improvements in mental health after anti-inflammatory treatment (p = 0.017), suggesting a subgroup of patients for whom initial poor mental health was inflammation-driven. Cross-lagged structural equation models indicated that reductions in diarrhea and abdominal pain preceded reductions in psychological symptoms over time (*β*=0.75, p <.0001), but not vice versa.

**Conclusions:** Baseline psychological distress is predictive of increased GI symptom severity and reduced mental HRQoL over time, suggesting early assessment of psychological symptoms may identify patients who may have worse disease trajectories. Abdominal pain predicted increased psychological distress, but not the other way around. Thus, intervening on abdominal pain may help prevent or reduce future psychological distress.

## Introduction

Ulcerative Colitis (UC) is an inflammatory bowel disease (IBD) characterized by mucosal inflammation of the rectum and colon. The most common symptom among UC patients is bloody diarrhea with mucus and urgency to evacuate.^1^ The disease course in UC is varying, with some patients having a mild disease course with few relapses and other patients having a severe disease course with frequent relapses leading to advanced therapy or surgery.^2^ Further, even in the absence of mucosal inflammation, approximately 25% patients may continue to experience irritable bowel syndrome(IBS)-like symptoms such as abdominal pain, bloating, and diarrhea.^3^

Given the clinical unpredictability of IBD, identifying factors that contribute to UC disease course may aid in the detection of early intervention targets that could potentially modulate disease outcomes. Prior research has found that biomarkers such as fecal calprotectin and serum IL-17A levels predict UC disease course in patients with new onset UC.^4,5^ In addition to biomarkers, psychological symptoms have been implicated as potential factors relevant to the clinical disease course. Indeed, up to one third and one quarter of IBD patients report symptoms of anxiety and depression, respectively,^6^ and several studies have demonstrated that baseline psychological symptoms are predictive of disease-related outcomes, including clinical relapses, escalation of therapy, hospitalization, emergency room visits, and surgery.^7-11^ Research suggests the relationship between IBD-related factors and psychological processes may be bidirectional,^8,12^ as disease status and IBD symptoms also predict the development of psychological symptoms.^8,12,13^ Further, both psychological symptoms and IBD-related factors (e.g., symptoms, disease status) have been found to influence health-related quality of life (HRQoL).^13,14^

While associations between disease-related outcomes, psychological symptoms, and HRQoL have been described in IBD, many studies are either cross-sectional^15^ or only evaluate baseline predictors of longitudinal outcomes^7-13^ as opposed to the dynamic relationship between these factors over time. Thus, there is limited knowledge elucidating the longitudinal evolution of UC symptoms and HRQoL, the biopsychosocial processes at onset determining this evolution, and the directionality of such effects.

To address this gap in the literature, the current study aimed to 1) study individual differences in longitudinal evolution of GI symptom levels (diarrhea, abdominal pain) and HRQoL (physical and mental aspects) by identifying subgroups based on these trajectories, 2) identify baseline predictors of subgroup allocation, and 3) disentangle the directionality of effects between GI symptom levels and level of psychological distress over time, while controlling for systemic inflammation levels.

## Methods

### Subjects

Ninety-eight patients with newly diagnosed UC based on clinical, endoscopic and histological findings (aged between 18-75 years) were included in the study. They were recruited from two outpatient gastroenterology clinics (Sahlgrenska University Hospital, Gothenburg, and Södra Älvsborgs Hospital, Borås) between April 2004 and June 2007. Patients were excluded if they were suffering from malignancy 5 years prior to inclusion, alcohol or drug abuse, significant heart, lung, kidney, neurological, or rheumatic disease, or major psychiatric disorders such as bipolar disorder, schizophrenia, or personality disorders (patients with depression or anxiety disorders were not excluded from participation). All patients provided written informed consent. The study was approved by the Regional Ethical Review Board at the University of Gothenburg.

Other results from this database have been reported before. More details about the patient sample and disease course can be found in these papers.^4,5,16-21^

### Study design

Biopsies from an inflamed area in the rectum/rectosigmoidal junction, blood samples, fecal samples, and self-report questionnaires were obtained at study inclusion (i.e. at diagnosis). Patients were then followed up at three months and then yearly for three years. An overview of the medication use at each time point can be found in Table 1. An overview of when the data / samples that are reported in this manuscript were collected can be found in Table 2. Since the predictive value of fecal calprotectin on disease course in this sample was already reported on earlier^4^ and since stool samples at disease onset were missing for 29 patients, data from the stool samples were not used in the current study.

**Table 1.** Medication use at each time point.

|  | Disease onset | 3-month FU | 1-year FU | 2-year FU | 3-year FU |
| --- | --- | --- | --- | --- | --- |
| Aminosalicylic acids | 95.9% | 81.6% | 76.6% | 81.5% | 70.65% |
| Corticosteroids | 53.1% | 15.3% | 9.6% | 8.7% | 3.3% |
| Thiopurines | 0% | 8.2% | 16.0% | 22.8% | 24.2% |

**Table 2.** Overview of data / samples per measurement moment.

|  | Disease onset | 3-month FU | 1-year FU | 2-year FU | 3-year FU |
| --- | --- | --- | --- | --- | --- |
| Questionnaires | X |  | X | X | X |
| Psychiatric interview |  | X | X |  |  |
| Mayo score | X |  | X | X | X |
| Endoscopy | X |  | X | X | X |
| Biopsies | X |  |  |  |  |
| Blood samples | X |  | X | X | X |

**Table 2.** Descriptive statistics of the variables at all measurement moments.xs.

|  | Theoretical range | Disease onset |  | 1-year follow-up |  | 2-year follow-up |  | 3-year follow-up |  |
| --- | --- | --- | --- | --- | --- | --- | --- | --- | --- |
|  |  | mean | SD | mean | SD | mean | SD | mean | SD |
| GSRS – diarrhea | 1 - 7 | 4.97 | 1.76 | 2.20 | 1.43 | 2.25 | 1.44 | 2.17 | 1.42 |
| GSRS – abdo pain | 1 - 7 | 2.46 | 1.18 | 1.81 | 0.88 | 1.79 | 0.87 | 1.62 | 0.79 |
| SF36 – PCS | 0 - 100 | 44.63 | 10.1 | 51.26 | 7.9 | 51.32 | 7.8 | 52.07 | 7.20 |
| SF36 – MCS | 0 - 100 | 40.11 | 12.8 | 46.00 | 12.0 | 48.11 | 10.4 | 47.15 | 10.2 |
| HADS total score | 0 - 28 | 11.63 | 4.72 | 9.98 | 3.79 | 10.02 | 4.02 | 9.48 | 3.86 |
| CRI | 60 - 240 | 173.3 | 25.4 | 173.1 | 25.5 | 170.4 | 32.8 | 175.2 | 27.0 |
| Mayo score | 0 - 12 | 7.58 | 2.40 | 1.17 | 2.05 | 0.96 | 1.87 | 1.07 | 2.13 |
| WBC |  | 8.18 | 3.13 | 7.24 | 2.05 | 7.07 | 2.27 | 6.76 | 2.28 |
| CRP |  | 21.56 | 30.3 | 5.89 | 4.8 | 5.05 | 3.2 | 5.48 | 5.8 |

### Measures

#### Self-report Questionnaires

- The *abdominal pain* and *diarrhea* subscales of the Gastrointestinal Symptom Rating Scale (GSRS)^22^ were used to measure self-reported GI symptom severity. Both scales range from 1 to 7 with higher scores representing higher discomfort due to pain / diarrhea.
- The MOS 36-item Short Form Health Survey (SF-36)^23^ was used to measure HRQoL. The physical (PCS) and mental (MCS) composite score were used to quantify mental and physical HRQoL. These measures have a theoretical range of 0 – 100 with higher scores indicating higher HRQoL, and 50 representing an average healthy adult according to normative values^24^.
- The Hospital Anxiety and Depression Scale (HADS)^25^ consists of 14 items on a Likert scale from 0-3 measuring depressive and anxiety symptoms. Given that the HADS does not consistently differentiate between anxiety and depression^26^, the total score of the HADS was used to measure general psychological distress.
- The total score of the Coping Resources Inventory (CRI)^27^ was used to measure coping. The CRI consists of 60 items on a 1 to 4 Likert scale, with higher score reflecting use of more effective coping resources.

#### Psychiatric interview

The Structured Clinical Interview for DSM-IV – Patient edition (SCID-P, DSM-IV)^28^, a structured diagnostic interview to assess diagnostic criteria for axis-I psychiatric disorders, was administered by a psychiatrist, trained in managing this diagnostic interview procedure, at two time points: 3-4 months after disease onset, when most patients were in remission, and at 1-year follow-up. For the purpose of this study, we extracted whether patients fulfilled the criteria for depressive disorder and for an anxiety disorder.

#### Mayo Score

The Mayo score for UC^29^ was used as a measure of disease severity. The Mayo scores combines self-report with clinical findings by calculating a sum score based on four observations: stool frequency, severity of rectal bleeding, disease activity based on endoscopic findings, and the physicians global assessments. A higher score indicates higher disease severity.

#### Inflammation markers

- Serum C-reactive protein (CRP) and white blood cell count (WBC) were used as measures of systemic inflammation.
- Expression of GATA, Tbet, FOX, IL-8, IL-6, IFN-γ, IL-13, IL-17A and RORC2 was quantified from the mucosal biopsies by RT-PCR
- Levels of the follow cytokines were measured in serum: IFN-γ, IL-17A, TNF-α, IL-10, IL-1β (Luminex, Flourokine MAP; R&D Systems)
- The following cytokines secreted by T-cells were measured in cell cultures of peripheral blood monocytes polyclonally stimulated with anti-CD3 and anti-CD28 for 24 hours: IL-10, TNF-α, IFN-γ, IL-13, IL-17 and IL-1β.

### Statistical analysis

The goal of the current study was to 1) identify subgroups based on longitudinal evolution of self-reported GI symptoms and HRQoL, 2) identify biopsychological risk factors of group membership at baseline and 3) investigate the directionality of the psychological functioning and GI symptoms relationships over time.

The main outcome variables were abdominal pain and diarrhea (GSRS) and mental and physical HRQoL (SF-36). The main predictor variables were psychological distress (HADS total score), coping resources (CRI), the Mayo score, and inflammation markers.

First, because of the large number of mucosal, serum, and T-cell secreted inflammation markers, principal component analysis with Varimax rotation was applied to the reduce the number of these markers (at baseline) into a smaller number of components, while still maintaining a high amount of explained variance. We carried out three separate principal component analyses, one for each category of inflammatory variables (mucosal, T-cell secreted, serum). In each analysis, the number of components was identified based on a Scree plot and the criterion of Eigenvalue>1.0.

Second, latent class growth analysis (LCGA)^30^ was used to identify subgroups in the evolution of self-reported symptom severity. LCGA is a data-driven analysis technique in which individuals are clustered together based on the combination of intercept and (linear and higher-order) slope of the individual trajectories of a variable measured over time. The number of clusters is decided based on fit using the Bayesian Information Criterion (BIC) such that the BIC of a solution with k+1 classes must be lower than the BIC of a solution with k classes, indicating a better fit of the k+1 cluster solution. Four separate LCGA analyses were performed using the SAS macro TRAJ^31^ on the following variables: GSRS-diarrhea, GSRS-abdominal pain, SF36-PCS, and SF36-MCS. Once the class solution for each variable was identified, risk factor analysis was performed using the same macro, to identify the extent to which variables measured at disease onset predicted class membership. Baseline risk factors included HADS total score, CRI total score, Mayo score, CRP, WBC, and the different factor scores representing serum, mucosal and T-cell secreted inflammation markers, as well as diarrhea and abdominal pain for the LCGA analyses on SF-36/HRQOL.

Third, cross-lagged panel models were built to investigate the directionality of the relationship between self-reported GI symptom severity and psychological functioning over time (see figure 1). More specifically, we investigated the extent to which psychological functioning at time T predicts changes in self-reported GI symptom severity at time T+1 (light grey dashed lines in figure 1) and vice versa (dark grey dotted lines in figure 1), while controlling for all auto-regressive coefficients (i.e. stabilities over time, the extent to which variable X at time T predicts variable X at time T+1, full black lines in figure 1) and within-time correlations between GI symptom severity and psychological functioning (thin grey dotted lines in figure 1). Additionally, we controlled for inflammation by adding paths from WBC at time T to GI symptom severity at time T+1 as well as within-time correlations between WBC and GI symptom severity and psychological functioning. WBC was chosen as a marker of inflammation in these analyses since this was the only marker that was not strongly zero-inflated during the follow-up measurement moments, at which many patients were in remission. Four separate cross-lagged panel models were constructed to investigate the relationship between 1) diarrhea and psychological distress (HADS total score), 2) abdominal pain and psychological distress, 3) diarrhea and coping resources and 4) abdominal pain and coping resources over time. While the strength of the autoregressive coefficients was not fixed over time, the strength of the cross-lagged paths was fixed over time in each direction since this provided better fit compared to freeing them.

**Figure 1.**
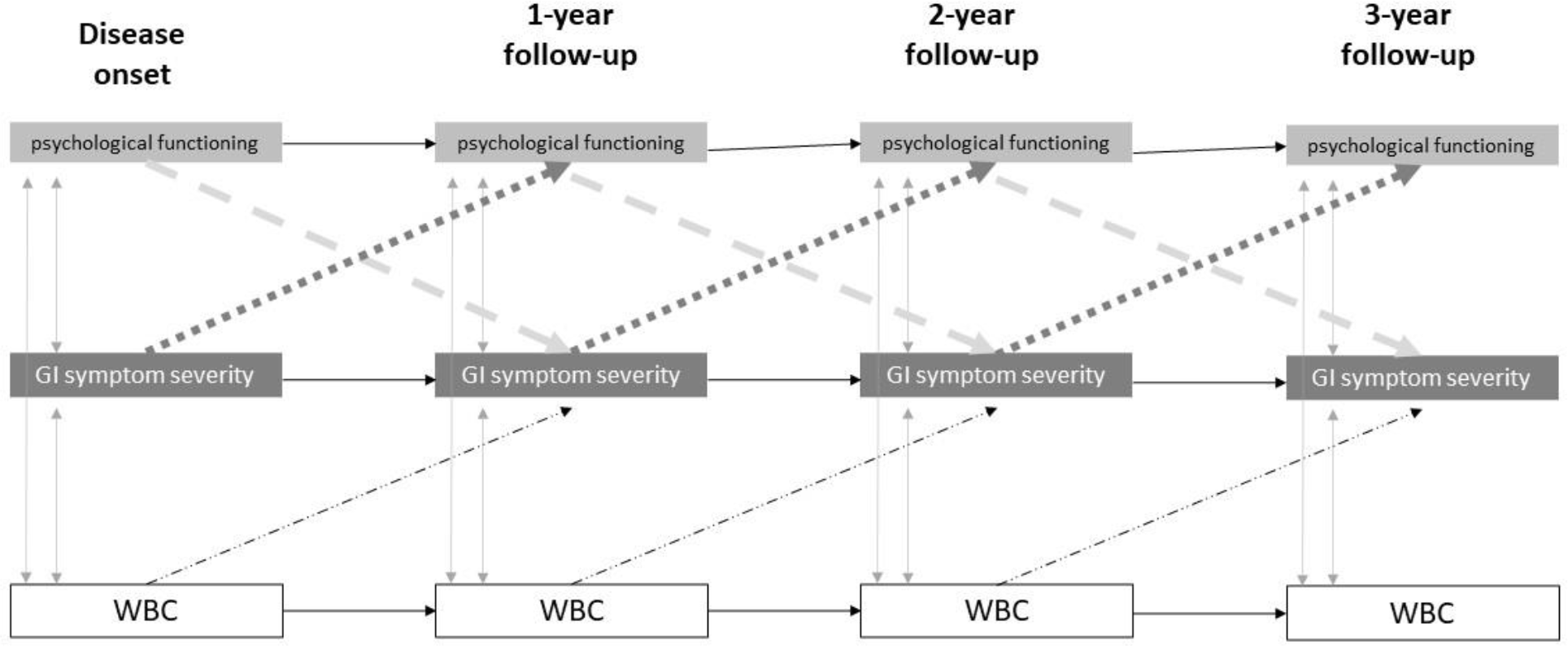
Example of the cross-lagged panel models in which the directionality of relationships between psychological functioning (HADS, CRI) and gastro-intestinal (GI) symptom severity over time was investigated, while controlling for white blood cell count (WBC). Paths of interest are the cross-lagged paths going from psychological functioning at time T to GI symptom severity at time T+1 (light grey dashed lines in figure 1) and vice versa (dark grey dotted lines in figure 1). This is investigated while controlling for all auto-regressive coefficients (i.e. stabilities over time, the extent to which variable X at time T predicts variable X at time T+1, full black lines) and within-time correlations between GI symptom severity and psychological functioning (thin grey dotted lines).

Variables that were not normally distributed were Box-Cox transformed before analysis. All analyses were performed with SAS 9.4 software (SAS Institute, Cary, NC, USA).

## Results

### Patient characteristics and descriptives

The study population consisted of 98 patients with new onset UC, 69% (*n* = 68) of whom were men. The mean age was 36 years (range 18–74). Colonic disease extent at the time of diagnosis was distributed as follows: proctitis 17% (*n* = 17), left-sided colitis 30% (*n* = 29) and extensive disease 53% (*n* = 52). Detailed results on the disease course in terms of evolution of inflammation and Mayo scores in this sample have been reported elsewhere.^16-21^

According to the SCID psychiatric interview, 11.8% of patients met criteria for a current depression and 15.1% of patients met criteria for an anxiety disorder at the first interview (3-4 months after disease onset), while 8.1% and 11.6% of patients fulfilled criteria for depression and anxiety respectively at follow-up 1. McNemar’s tests indicated that there was no significant difference in prevalence of anxiety (χ^2^ = 1.33, p = 0.25) or depression (χ^2^ = 1.00, p = 0.32) between the 2 time points. Descriptive statistics of questionnaire scores, Mayo scores, WBC and CRP levels can be found in Table 2.

### Principal Component Analysis

For the mucosal inflammatory markers, principal component analysis produced three components: Mucosal Component 1 (4 items, Eigenvalue = 3.18) comprised of GATA, Tbet, FOX, and IL-8; Mucosal Component 2 (2 items, Eigenvalue = 1.72) comprised of IL-6 and IFN-γ; Mucosal Component 3 (3 items, Eigenvalue = 1.35) comprised of RORC2, IL-17A, and IL-13.

For the T-cell secreted cytokines, five of the six variables (IL-10, TNF-α, IFN-γ, IL-13, IL-17A, IL-1β) loaded onto one component named T-cells Cytokines Component 1 (Eigenvalue = 2.62). The remaining T-cell secreted cytokine was IL-1β, which was entered into the subsequent models on its own.

For the serum inflammatory markers, the analyses produced two components: Serum Component 1 (3 items, Eigenvalue = 1.77), comprised of IFN-γ, IL-17A, and TNF-α; Serum Component 2 (2 items, Eigenvalue = 1.10), comprised of IL-10 and IL-1β.

Please see Supplement for more information (i.e., factor pattern and loadings) for the principal component analysis.

### Latent class growth analysis and risk factor analysis

Subgroups in longitudinal trajectories based on intercept and slope were investigated for 4 variables (GSRS – diarrhea, GSRS – abdominal pain, SF36 – PCS and SF36 – MCS). The estimated trajectories are depicted in Figure 2, while the LCGA solutions are depicted in tables 3-6. For interpretation of the slopes in the tables, note that 1 unit of the “time” variable equals 1 month.

**Table 3.** Solution for the latent class growth analysis on GSRS – diarrhea. Slopes with an asterisk (*) are significantly different from 0 at p < 0.05. Numbers with the same superscript letter (a, b, c) within the same column are not significantly (p > 0.05) different from one another.

|  | % of sample | Intercept | Linear slope | Quadratic slope | Cubic slope |
| --- | --- | --- | --- | --- | --- |
| Class 1<br>*Low diarrhea class* | 35.4% | 3.05405<sup>a</sup> | -0.20164<sup>*a</sup> | 0.00778<sup>a</sup> | -0.00009<sup>a</sup> |
| Class 2<br>*Diarrhea improvement class* | 40.7% | 6.35472<sup>b</sup> | -0.68139<sup>*b</sup> | 0.03068<sup>*b</sup> | -0.00042<sup>*b</sup> |
| Class 3<br>*Residual diarrhea class* | 23.9% | 5.98971<sup>b</sup> | -0.42625<sup>*a</sup> | 0.02385<sup>*ab</sup> | -0.00039<sup>*ab</sup> |

**Table 4.** Solution for the latent class growth analysis on GSRS – abdominal pain. Slopes with an asterisk (*) are significantly different from 0 at p < 0.05. Numbers with the same superscript letter (a, b, c) within the same column are not significantly (p > 0.05) different from one another.

|  | % of sample | Intercept | Linear slope | Quadratic slope |
| --- | --- | --- | --- | --- |
| Class 1<br>*Low abdominal pain class* | 81.4% | 2.11165<sup>a</sup> | -0.04792<sup>*a</sup> | 0.00081<sup>*a</sup> |
| Class 2<br>*Medium abdominal pain class* | 18.6% | 3.74089<sup>b</sup> | -0.06226<sup>*a</sup> | 0.00087<sup>a</sup> |

**Table 5.** Solution for the latent class growth analysis on SF36 - PCS. Slopes with an asterisk (*) are significantly different from 0 at p < 0.05. Numbers with the same superscript letter (a, b, c) within the same column are not significantly (p > 0.05) different from one another.

|  | % of sample | Intercept | Linear slope | Quadratic slope | Cubic slope |
| --- | --- | --- | --- | --- | --- |
| Class 1<br>*Low physical HRQoL class* | 10.6% | 34,89325<sup>a</sup> | 0,6771<sup>a</sup> | -0,05566<sup>ab</sup> | 0,00111<sup>ab</sup> |
| Class 2<br>*Physical HRQoL improvement class* | 27.4% | 34,65773<sup>a</sup> | 2,60326<sup>*b</sup> | -0,12154<sup>*a</sup> | 0,00173<sup>*a</sup> |
| Class 3<br>*High physical HRQoL class* | 62.0% | 50,78158<sup>b</sup> | 0,39459<sup>a</sup> | -0,01432<sup>b</sup> | 0,00017<sup>b</sup> |

**Table 6.** Solution for the latent class growth analysis on SF36 - MCS. Slopes with an asterisk (*) are significantly different from 0 at p < 0.05. Numbers with the same superscript letter (a, b, c) within the same column are not significantly (p > 0.05) different from one another.

|  | % of sample | Intercept | Linear slope | Quadratic slope |
| --- | --- | --- | --- | --- |
| Class 1<br>*Low mental HRQoL class* | 8.5% | 21,40614<sup>a</sup> | 0,01629<sup>a</sup> | 0,00499<sup>a</sup> |
| Class 2<br>*Mental HRQoL improvement class* | 37.4% | 31,70478<sup>b</sup> | 1,11308<sup>*b</sup> | -0,02173<sup>*b</sup> |
| Class 3<br>*High mental HRQoL class* | 54.1% | 48,67825<sup>c</sup> | 0,38176<sup>*a</sup> | -0,0077<sup>a</sup> |

**Figure 2.**
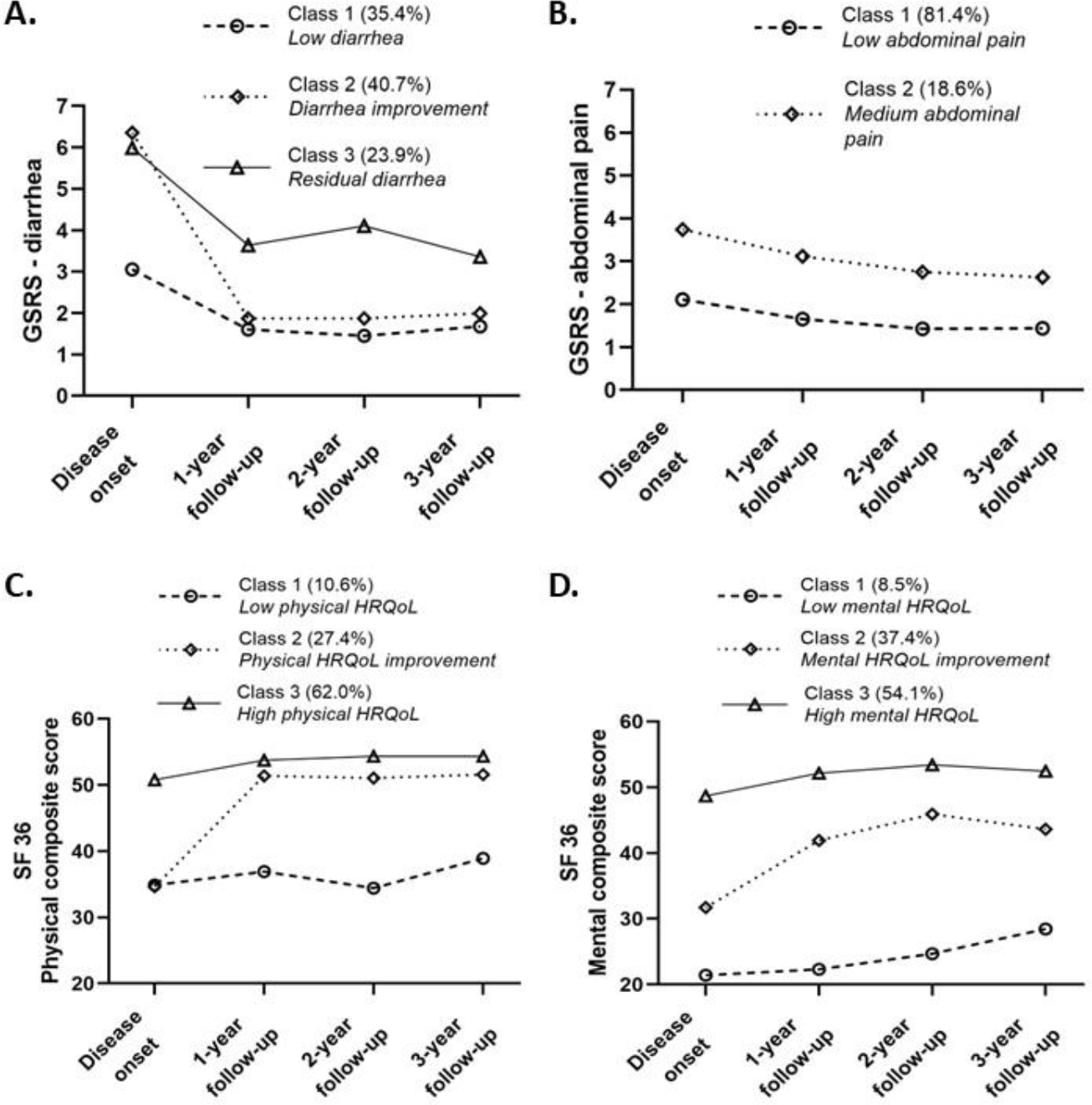
Estimated symptom trajectories based on intercepts and slopes of the LCGA solution for A. GSRS – diarrhea, B. GSRS – abdominal pain, C. SF36 – PCS, and D. SF36 – MCS.

#### 1. GSRS – diarrhea

Table 3 shows the LCGA solution for subgroups in trajectories of self-reported diarrhea severity as measured with the GSRS. As can be seen in Figure 2A, 3 distinct classes of diarrhea trajectory were identified. Both Class 2 and Class 3 had high diarrhea scores at disease onset, but while patients in Class 2 had large reductions in diarrhea following anti-inflammatory treatment *(= “diarrhea improvers class”*, 40.7% of sample), patients in class 3 were still burdened by significant diarrhea symptoms at the follow-up moments *(= “residual diarrhea class*, 23.9% of sample). Patients in class 1 had lower levels of diarrhea at disease onset, which kept decreasing during follow-up *(“low diarrhea class”*, 35.4% of sample).

Risk factor analysis was used to investigate whether baseline (= at disease onset) levels of psychological functioning, illness severity, and inflammation influenced the probability of being allocated to one class compared to the others. Patients with higher total HADS scores (= higher levels of psychological distress) at disease onset had a lower chance of being allocated to the low diarrhea class compared to the residual diarrhea class (t = -1.98, p = 0.049). Patients with higher Mayo scores (= higher clinical disease severity) at disease onset had lower chances of being allocated to the low diarrhea class compared to the diarrhea improvement (t = -2.78, p = 0.006) and the residual diarrhea (t = -2.41, p = 0.017) classes. Finally, patients with higher levels of CRP at disease onset had a lower chance of being allocated to the low diarrhea class compared to the residual diarrhea class (t = -2.62, p = 0.009). No associations were found between baseline CRI scores, mucosal, serum, and T-cell secreted inflammation markers, and WBC on the one hand and diarrhea trajectory subgroup allocation on the other hand.

#### 2. GSRS – abdominal pain

Table 4 shows the LCGA solution for subgroups in trajectories of self-reported abdominal pain. Two distinct classes of abdominal pain trajectory were identified (Figure 2B), that could only be distinguished by differences in intercept but not changes in rate of change. The large majority of patients had stable low levels of abdominal pain (class 1, *“low abdominal pain class”*, 81.1%) throughout the study, while a minority of patients had stable medium levels of abdominal pain throughout the study *(“medium abdominal pain class”*, 18.6%). Risk factor analysis indicated that patients with higher levels of psychological distress at disease onset had lower chance of being allocated to the low abdominal pain class compared to the medium abdominal pain class (t = -2.63, p = 0.009). None of the other predictors were associated with subgroup allocation.

#### 3. SF36 – Physical composite score

Table 5 shows the LCGA solution for subgroups in trajectories of physical HRQoL as measured with the physical composite score of the SF36. Three distinct trajectories were identified (Figure 2C). The majority of patients had high physical HRQoL at disease onset - a score of 50 represents an average healthy adult – and this did not change throughout the study (class 3, “*High physical HRQoL class”*, 62.0% of sample). A small minority of patients had stable low levels of physical HRQoL (class 1, *“Low physical HRQoL class”*, 10.6% of sample). A third group of patients had equally low physical HRQoL at disease onset but improved to a healthy level of around 50 after anti-inflammatory treatment (class 2, *“Physical HRQoL improvement class”*, 27.4% of sample). Risk factor analysis indicated that patients with higher CRP levels at disease onset had a lower chance of being allocated to the high physical HRQoL class compared to the physical HRQoL improvement class (p = -2.83, p = 0.005). Moreover, patients who had higher levels of diarrhea and abdominal pain at disease onset had a lower chance of belonging to the high physical HRQoL class compared to the low physical HRQoL class (diarrhea: t = -2.75, p = 0.006; abdominal pain: t = -3.05, p = 0.002) and the physical HRQoL improvement class (diarrhea: t = -2.51, p = 0.012, abdominal pain: t = -3.14, p = 0.002)

#### 4. SF36 – Mental composite score

Table 5 shows the LCGA solution for subgroups in trajectories of mental HRQoL as measured with the mental composite score of the SF36. Three distinct trajectories were identified (Figure 2D). More than half of the patients had high mental HRQoL at disease onset and little change throughout the study (class 3, “*high mental HRQoL class”*, 54.1% of sample). A small minority of patients (8.5%) had very low mental HRQoL at disease onset which did not improve after treatment (Class 1, *low mental HRQoL class)*. Class 2 consisted of 37.4% of patients and was characterized by rather low mental HRQoL at disease onset with great improvements throughout the study (“*mental HRQoL improvement class”)*.

Unsurprisingly, patients with higher levels of depression at disease onset had a lower chance of being classified as high mental HRQoL relative to low mental HRQoL (t = -3.80, p < 0.001) and mental HRQoL improvement (t = -3.42, p < 0.001). Similarly, patients with higher levels of coping resources at disease onset had a higher chance of being classified as high mental HRQoL relative to low mental HRQoL (t = 2.11, p = 0.036) and mental HRQoL improvement (t = 2.54, p = 0.011). Patients who had higher levels of diarrhea and abdominal pain at disease onset had a lower chance of belonging to the high mental HRQoL class compared to the low mental HRQoL class (diarrhea: t = -3.63, p < 0.001; abdominal pain: t = -2.94, p = 0.003) and the physical HRQoL improvement class (diarrhea: t = -3.12, p = 0.002, abdominal pain: t = -3.18, p = 0.002). Interestingly, patients with higher levels of CRP at disease onset had a higher chance of being allocated to the mental HRQoL improvement class compared to the low mental HRQoL class (t = 2.40, p = 0.017).

### Cross-lagged panel analysis

To disentangle directionality of the longitudinal relationship between psychological functioning and GI symptom severity, four separate cross-lagged models were investigated.

### Psychological distress (HADS) and abdominal pain (Figure 3)

Abdominal pain at time T positively predicted changes in psychological distress at time T+1 (standardized β = 0.165 – 0.238, all p’s < 0.001), indicating that patients with more abdominal pain at time T experienced greater increases in psychological distress from time T to time T+1. However, psychological distress at time T did not predict changes in diarrhea at time T+1 (standardized β = 0.049 – 0.060, p = 0.35 - 0.36).

**Figure 3.**
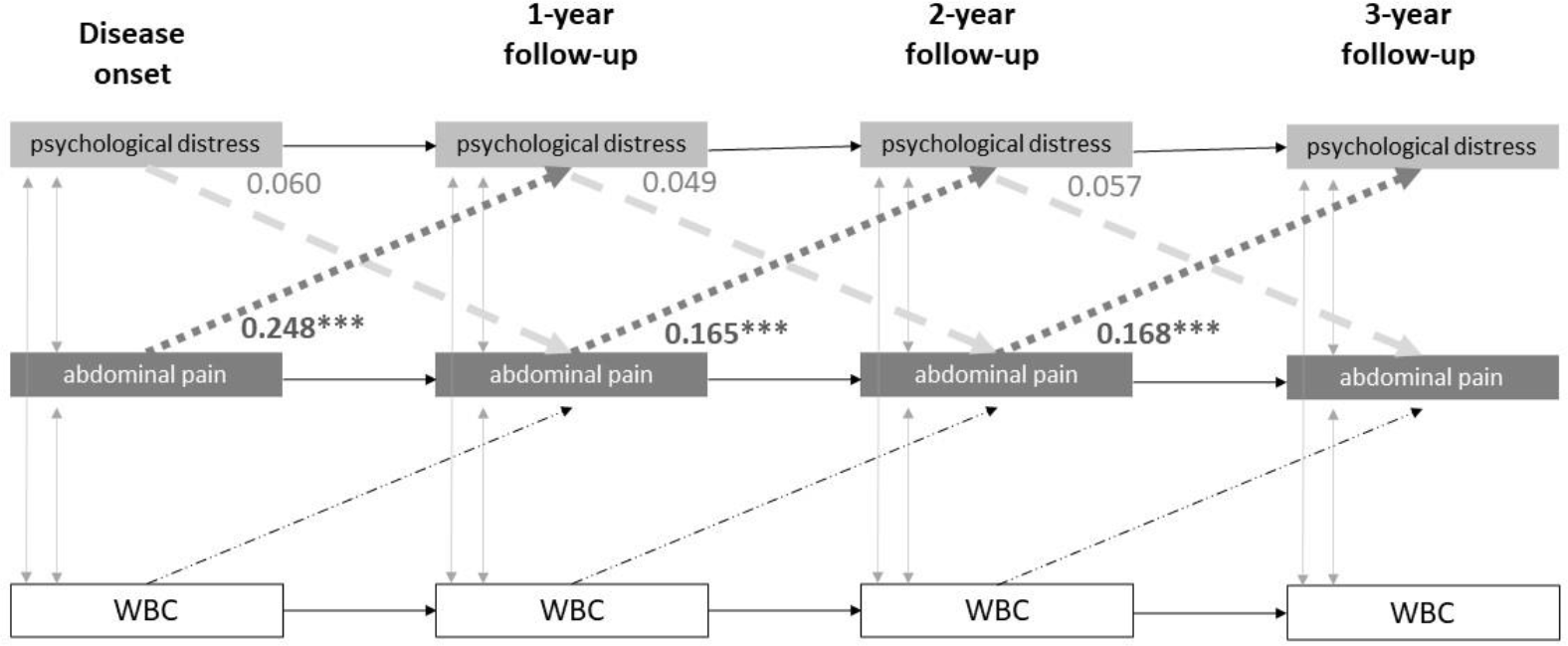
Cross-lagged panel model investigating dynamic relationship between psychological distress and abdominal pain over time. Numbers represent standardized path coefficients, ***p < 0.001.

#### Psychological distress (HADS) and diarrhea (Figure 4)

The cross-lagged paths going from diarrhea to psychological distress were not significant (standardized β = -0.003, p = 0.96), nor vice versa (standardized β = 0.037 – 0.049, p = 0.34 - 0.35)

**Figure 4.**
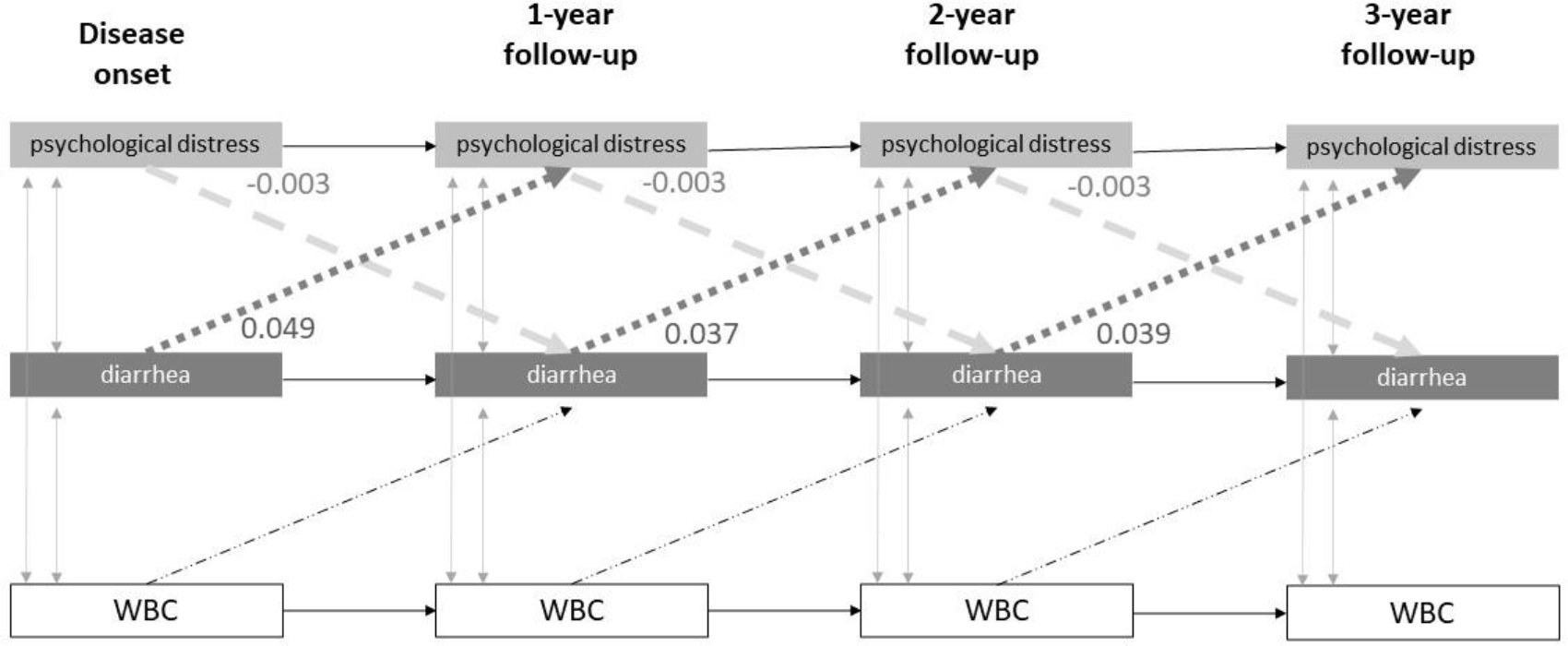
Cross-lagged panel model investigating dynamic relationship between psychological distress and diarrhea over time. Numbers represent standardized path coefficients.

#### Coping resources (CRI) and abdominal pain (Figure 5)

Abdominal pain at time X negatively predicted changes in coping resources at time T+1 (standardized β = -0.178 – -0.310, all p’s < 0.001), indicating that patients with more abdominal pain at time T experienced smaller increases in coping resources from time T to time T+1. However, coping resources at time T did not predict changes in diarrhea at time T+1 (standardized β = -0.036 – -0.053, p = 0.41).

**Figure 5.**
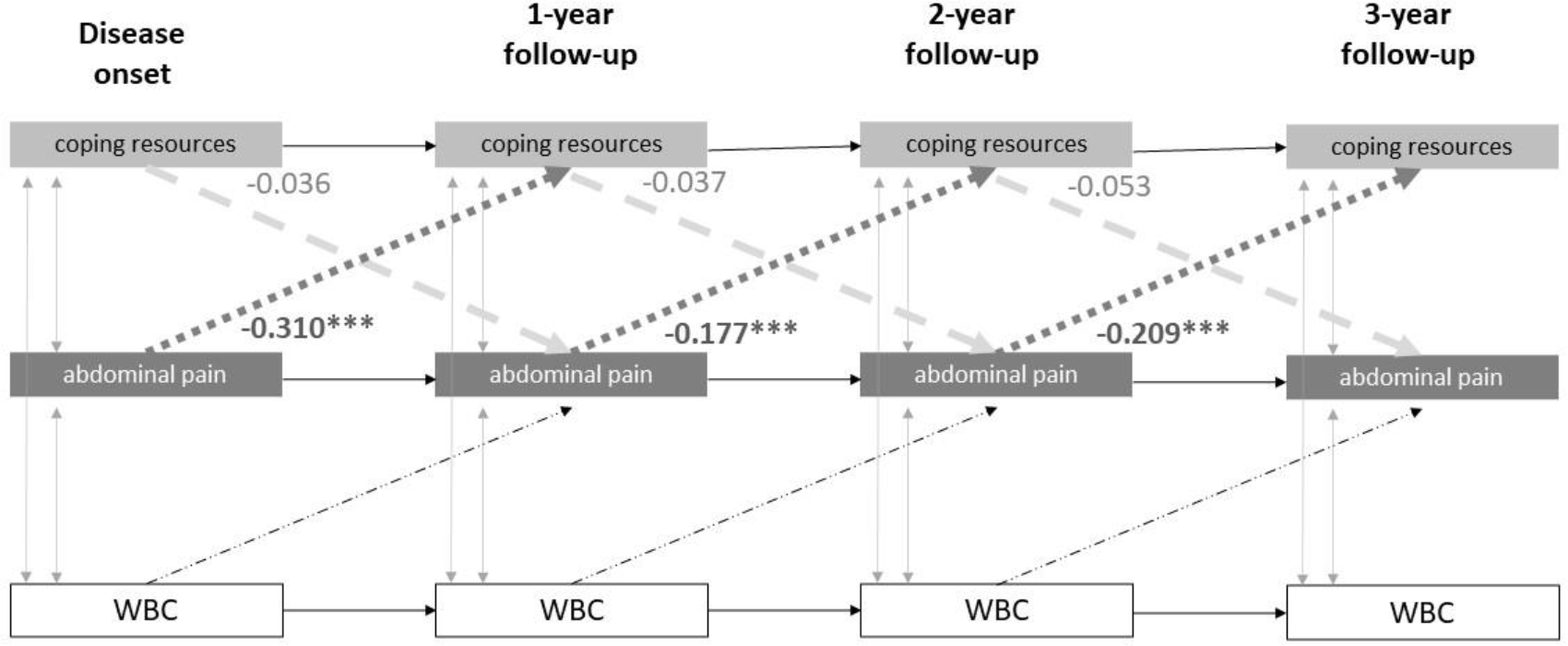
Cross-lagged panel model investigating dynamic relationship between coping resources and abdominal pain over time. Numbers represent standardized path coefficients, ***p < 0.001.

#### Coping resources (CRI) and diarrhea (Figure 6)

Diarrhea at time X negatively predicted changes in coping resources at time T+1 (standardized β = -0.096 – -0.155, p = 0.011 - 0.012), indicating that patients with more diarrhea at time T experienced smaller increases in coping resources from time T to time T+1. However, coping resources at time T did not predict changes in diarrhea at time T+1 (standardized β = 0.020 – 0.021, p = 0.68).

**Figure 6.**
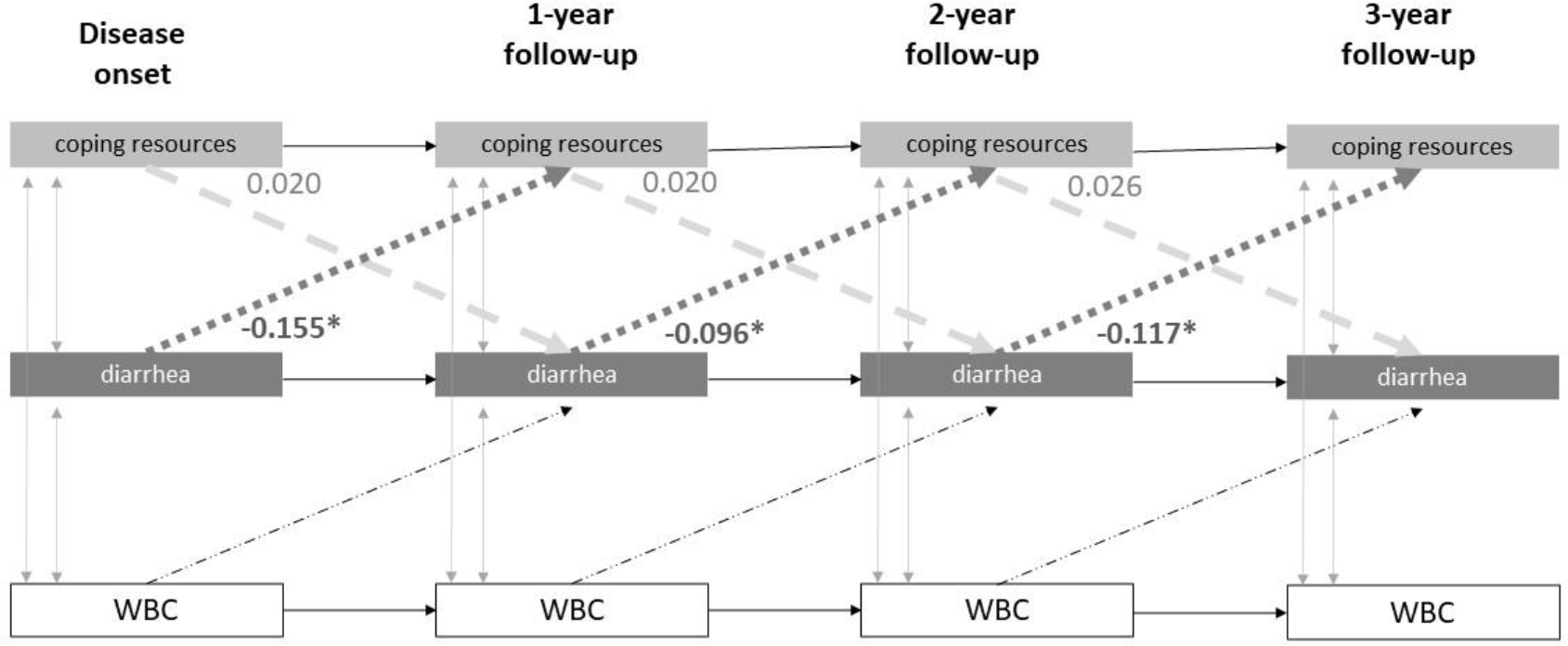
Cross-lagged panel model investigating dynamic relationship between coping resources and diarrhea over time. Numbers represent standardized path coefficients, *0.01 < p < 0.05.

## Discussion

The goal of this study was to investigate individual differences in predictors of self-reported GI symptom severity and HRQoL trajectories in a longitudinal study following up patients with newly onset UC. We used LCGA to identify subgroups in GI symptom / HRQoL trajectories over time. For diarrhea and physical and mental HRQoL, we consistently found three subgroups: one with stable low levels of symptom severity (high HRQoL, respectively), one with stable high levels of symptom severity (low HRQoL, respectively), and one with initial high levels of symptom severity (low levels of high HRQoL) showing great improvements at follow-up (after anti-inflammatory treatment). Risk factor analysis was used to investigate whether levels of psychological functioning and inflammation at disease onset could predict subgroup allocation. Although predictors that would be able to distinguish between improvers and non-improvers in patients with high initial levels of symptom burden would have been most informative, the included predictors were mainly successful in distinguishing groups that already had different levels of symptom burden at disease onset. Thus, while psychological processes are related to disease trajectory, this seemed to be mostly a baseline effect. Moreover, cross-lagged analysis revealed that in this study, symptom severity predicted changes in psychological functioning over time, and not vice versa.

Our study identified baseline prevalence rates for anxiety (15.5%) and depression (11.8%) that remained stable over time, which were lower than those previously reported in the literature.^6,32^ A recent systematic review and meta-analysis found a pooled-prevalence of 32.1% for anxiety and 25.2% for depression.^6^ One potential reason for the lower prevalence rates in our study may be due to differences in assessment methods. Most studies use self-report questionnaires, which evaluate the presence of psychological symptoms. Our study used a validated structural interview to assess diagnostic criteria for depression and anxiety that was administered by trained mental health clinicians (i.e., psychiatrists), which likely resulted in more conservative and accurate representation of psychiatric comorbidity. Further, many studies report the combined Crohn’s disease (CD) and UC prevalence, but when broken down by disease type, there is evidence for higher rates of anxiety and depression in CD compared to UC.^6^

When evaluating the longitudinal evolution of IBD symptoms, our study found that patients with higher psychological distress at baseline had an increased probability of maintaining higher levels of diarrhea and abdominal pain over time. These findings are consistent with recent research suggesting a relationship between baseline psychological symptoms and worse IBD-related outcomes at follow up.^8-10,12^ Our study adds to this literature by demonstrating the role of baseline psychological symptoms on longitudinal IBD *symptoms*, as opposed to other IBD-related outcomes (i.e., healthcare utilization). It must be noted, though, that psychological distress at disease onset did not predict which patients amongst those with initial high levels of IBD symptoms would improve and which would not; rather, lower psychological distress was related to lower IBD symptoms at disease onset, which was then maintained over time.

Specific to abdominal pain, we found two groups of disease trajectories – (1) low pain at baseline that remained low over time and (2) high pain at baseline that remained high over time. Interestingly, there were no differences in baseline inflammation, neither locally nor systemically, between the two groups. Psychological distress was the only baseline factor significantly different between the two groups, with higher psychological distress predicting membership in the group with more severe abdominal pain. One potential explanation for our findings is that those with maintained severe abdominal pain and higher psychological distress scores developed visceral hypersensitivity in the context of their IBD. Indeed, visceral hypersensitivity (evaluated via rectal balloon distention), along with psychological factors and female gender, is associated with the presence of GI symptoms, including abdominal pain, in UC patients in remission.^33^

Cross-lagged panel models evaluated the longitudinal relationship between abdominal pain and psychological distress, allowing us to disentangle the directionality of effects over time. We found that abdominal pain predicts psychological symptoms, and not the other way around, indicating that treating abdominal pain may help to prevent or reduce subsequent psychological distress. Treatments such as neuromodulators and brain-gut behavior therapies (e.g., cognitive-behavioral therapy (CBT), gut-directed hypnotherapy), have been shown to reduce abdominal pain in patients with IBD.^34^ A potential next step is to identify mediators of the relationship between abdominal pain and subsequent psychological distress, which could inform future treatment targets for brain-gut behavior therapy. One potential mechanism is pain catastrophizing, which has been found to mediate the relationship between abdominal pain and anxiety symptoms in youth (11-18 years old) with IBD.^35^

The finding that changes in symptom burden consistently precede changes in psychological distress in IBD is inconsistent with findings from similar studies in functional syndrome patients. For example, Van Oudenhove et al.^36^ report that levels of depression, stress, and anxiety at Time T predict changes in fatigue levels at Time T+1 in a group of patients with chronic fatigue syndrome following CBT, but not the other way around. However, an earlier study investigating predictors of treatment response in fibromyalgia patients following multidisciplinary treatment reported that reductions in pain-related disability and physical functioning preceded reductions in anxiety, depression, and coping^37^. One explanation for this is that a bidirectional relationship is possible, but that the first change will take place in the variable / component that is targeted directly by the specific treatment (e.g. anxiety and depression are targeted by CBT, while IBD symptoms are targeted by anti-inflammatory treatment).

For the LCGA models with diarrhea as an outcome, we were not able to distinguish between the groups with more severe diarrhea symptoms at baseline who improved and those who remained high based on any physiological or psychological baseline factors. In addition, there was no significant association between diarrhea symptoms and psychological distress in either direction, when evaluated in the cross-lagged panel models. One potential reason for these findings may be that the association between psychological symptoms and diarrhea is not as strong compared to abdominal pain. One study comparing psychological symptoms between patients with IBS and those with functional diarrhea and functional constipation found that IBS was associated with higher anxiety and depression scores^38^, suggesting that the presence of abdominal pain is linked to more psychological symptoms compared to other symptoms (i.e., diarrhea).

In addition to IBD symptoms, we also evaluated the longitudinal evolution of mental and physical HRQoL. Overall, patients with lower levels of baseline diarrhea and abdominal pain were more likely to be in groups with higher mental and physical HRQoL. Our findings align with two recent studies that found an association between baseline IBS-type symptoms and reduced HRQoL (also using the SF-36) at follow up in IBD.^13,39^ In mental HRQoL specifically, we found two groups with low levels of mental HRQoL at baseline but different trajectories (stable low vs. improvement) over time. When evaluating of trajectory group membership, the only significant effect was that individuals with high CRP at disease onset were more likely to belong to the mental HRQoL improvement class compared to the stable low mental HRQoL class. These findings suggest that poor mental HRQoL may be inflammatory-driven in the former subset of patients, as scores improve over time when inflammation is treated. It has been previously demonstrated that baseline disease activity is associated with elevations in psychological symptom reporting over time.^8,12^ However, research disentangling the relationship between inflammation and psychological reporting over time is needed to confirm.

Finally, prior research has identified associations between coping and IBD-related outcomes, with one study reporting associations between coping strategies and pain levels,^40^ and another study demonstrating an association between baseline maladaptive coping and increased IBD symptoms at 6-month follow up.^41^ We expanded on this literature by evaluating the causal relationships between IBD symptoms and coping resources over time. Our findings indicate that both abdominal pain and diarrhea precede reductions in coping resources, and not the other way around. Just as in the analysis with abdominal pain and psychological distress, findings underscore the importance treating IBD symptoms early to potentially prevent reductions in coping resources over time. As with psychological distress, brain-gut behavioral therapies may be useful to address maladaptive coping, as interventions can simultaneously target IBD-related symptoms and coping skills.^42^

Our study has several limitations that should be considered. First, the sample was recruited prior to the widespread use of biologic therapies. Thus, the disease trajectories found in our study may be different from current disease trajectories. Nevertheless, there is evidence of IBS-type symptom overlap, similar to what was found in our study, in patients treated with biologic therapies.^3^ Therefore, findings may still be relevant to patients with continued abdominal pain/diarrhea despite being in clinical remission with biologic therapy. For HRQoL, we found that a high number of people already reported high scores on the SF-36, particularly for the physical component. The physical component of the SF-36 is mostly assessing HRQoL-related difficulties due to musculoskeletal concerns (i.e., difficulty going up the stairs) and may not capture aspects of HRQoL affected by IBD. Thus, future research should include questionnaires assessing IBD-specific quality of life, such as the Inflammatory Bowel Disease Questionnaires (IBDQ).^43^

## Data Availability

All data produced in the present study are available upon reasonable request to the authors.

## Notes

**Grant Support:** Livia Guadagnoli is a postdoctoral research fellow of the Research Foundation Flanders (FWO, 12A7822N). Lukas Van Oudenhove is a research professor of the KU Leuven Special Research Fund.

### Competing Interest Statement

The authors have declared no competing interest.

### Funding Statement

Livia Guadagnoli is a postdoctoral research fellow of the Research Foundation Flanders (FWO, 12A7822N). Lukas Van Oudenhove is a research professor of the KU Leuven Special Research Fund.

### Author Declarations

The Regional Ethical Review Board of the University of Gothenburg gave ethical approval for this work.

