## Supplement for "Predictors of symptom trajectory in newly diagnosed ulcerative colitis: a 3-year follow-up cohort study"

Supplementary Table 1. Principal component analysis for mucosal inflammatory markers

| Rotated Factor Pattern | | | | | | | |
|---|---|---|---|---|---|---|---|
| | | Factor1 | | Factor2 | | Factor3 | |
| GATA_muc | GATA_muc | 96 | * | -3 | | 10 | |
| Tbet_muc | Tbet_muc | 94 | * | 13 | | 7 | |
| FOX_muc | FOX_muc | 85 | * | -14 | | 2 | |
| IL_8_muc | IL_8_muc | 78 | * | 22 | | -3 | |
| IL_6_muc | IL_6_muc | 7 | | 87 | * | -12 | |
| IFN_g_muc | IFN_g_muc | 7 | | 82 | * | 12 | |
| RORC2_muc | RORC2_muc | 2 | | 3 | | 83 | * |
| IL_17_muc | IL_17_muc | -9 | | 40 | | 63 | * |
| IL_13_muc | IL_13_muc | -13 | | 20 | | -48 | * |
| Printed values are multiplied by 100 and rounded to the nearest integer. Values greater than 0.4 are flagged by an '*'. | | | | | | | |

| Variance Explained by Each Factor | | |
|---|---|---|
| Factor1 | Factor2 | Factor3 |
| 3.1800497 | 1.7180195 | 1.3465209 |

Supplementary Table 2: Principal component analysis for T-cell derived inflammatory markers

| Factor Pattern | | | |
|---|---|---|---|
| | | Factor1 | |
| IL_10_supern | IL_10_supern | 76 | * |
| TNF_a_supern | TNF_a_supern | 75 | * |
| IFN_g_supern | IFN_g_supern | 74 | * |
| IL_13_supern | IL_13_supern | 70 | * |
| IL_17_supern | IL_17_supern | 65 | * |
| IL_1b_supern | IL_1b_supern | 13 | |
| Printed values are multiplied by 100 and rounded to the nearest integer. Values greater than 0.4 are flagged by an '*'. | | | |

| Variance Explained by Each Factor |
|---|
| Factor1 |
| **2.6201800** |

Supplementary Table 3: Principal component analysis for serum inflammatory markers

| Rotated Factor Pattern | | | | | |
|---|---|---|---|---|---|
| | | Factor 1 | | Factor 2 | |
| IFN_g_serum | IFN_g_serum | 82 | * | 14 | |
| IL_17_serum | IL_17_serum | 76 | * | -3 | |
| TNF_a_serum | TNF_a_serum | 61 | * | -40 | * |
| IL_10_serum | IL_10_serum | 25 | | 75 | * |
| IL_1b_serum | IL_1b_serum | 23 | | -61 | * |
| Printed values are multiplied by 100 and rounded to the nearest integer. Values greater than 0.4 are flagged by an '*'. | | | | | |

| Variance Explained by Each Factor | |
|---|---|
| Factor1 | Factor2 |
| 1.7491140 | 1.1135571 |
